# The effects of the first national lockdown in England on geographical inequalities in the evolution of COVID-19 case rates: An ecological study

**DOI:** 10.1101/2021.11.09.21266122

**Authors:** Claire E. Welsh, Viviana Albani, Fiona E. Matthews, Clare Bambra

**Author notes:** Corresponding author, Room 2.38 Biomedical Research Building, Campus for Aging and Vitality, Newcastle University, Newcastle, NE4 5PL. **Authorship** CW, CB and FM designed the study. CW completed all analyses with input from FM and CB. CW, VA, FM and CB all contributed to drafting the manuscript. The corresponding author attests that all listed authors meet authorship criteria and that no others meeting the criteria have been omitted. CW is guarantor of the analysis. The Corresponding Author has the right to grant on behalf of all authors and does grant on behalf of all authors, a worldwide licence to the Publishers and its licensees in perpetuity, in all forms, formats and media (whether known now or created in the future), to i) publish, reproduce, distribute, display and store the Contribution, ii) translate the Contribution into other languages, create adaptations, reprints, include within collections and create summaries, extracts and/or, abstracts of the Contribution, iii) create any other derivative work(s) based on the Contribution, iv) to exploit all subsidiary rights in the Contribution, v) the inclusion of electronic links from the Contribution to third party material where-ever it may be located; and, vi) licence any third party to do any or all of the above. **Data Sharing Statement** All data used are publicly freely available through the GOV.UK and the ONS. Code used in the analyses is available upon request. **Ethical Approval** This study was approved by the Newcastle University Ethics Committee (Ref: 7543/2020). **Transparency declaration** The lead author* affirms that this manuscript is an honest, accurate, and transparent account of the study being reported; that no important aspects of the study have been omitted; and that any discrepancies from the study as planned (and, if relevant, registered) have been explained. The manuscript’s guarantor.

## Abstract

**Background:** Socio-economic inequalities in COVID-19 case rates have been noted worldwide. Previous studieshave compared case rates over set phases. There has been no analysis of how inequalities in cases changed overtime and were shaped by national mitigation strategies (e.g. lock downs). This paper provides the first analysis of the evolution of area-level inequalities in COVID-19 cases by deprivation levels in the first wave of the pandemic (January to July 2020) in England – with a focus on the effects of the first national lockdown (March – July 2020).

**Methods:** Weekly case rates per Middle Super Output Area (MSOA, n=4412) in England from 2020-03-15 to 2020-07-04 were obtained, and characteristics of local epidemics were calculated, e.g. the highest case rate per area. Simple linear and logistic regression analyses were employed to assess the association of these metrics with index of multiple deprivation (IMD). Local authority-level (n=309) cases were used similarly in a sensitivity analysis, as these data were available daily and extended further back in time. The impact of lockdown was assessed by comparing the cumulative case rate in the most deprived 20% of MSOAs to the least deprived 20%, for the periods before the lockdown, and by the end of lockdown.

**Findings:** Less deprived areas began recording COVID-19 cases earlier than more deprived areas and were more likely to have peaked by March 2020. More deprived areas’ case rates grew faster and peaked higher than less deprived areas. During the first national lockdown in the UK, the relative excess in case rates in the most deprived areas increased to 130% of that of the least deprived ones.

**Interpretation:** The pattern of disease spread in England confirm the hypothesis that initial cases of a novel infectious disease are likely to occur in more affluent communities, but more deprived areas will overtake them once national mitigation strategies begin, and bear the brunt of the total case load. The strict first national lockdown served to increase case rate inequalities in England.

**Funding:** This work was supported by a grant from The Health Foundation (Ref: 2211473), who took no part in the design, analysis or writing of this study.

**Research in Context:** 

**Evidence before this study:** The magnitude and distribution of deprivation-related inequalities in COVID-19 cases have been reported for England and many other countries, however, none have yet investigated the initial evolution of these inequalities, nor the effects of the first national lockdown.

**Added value of this study:** We leverage the benefits of two separate datasets of COVID-19 case counts to investigate the initiation and evolution in inequalities in disease burden by deprivation. We found that cases were first recorded in less deprived areas before rising faster in more deprived areas. The first national lockdown led to an increase in these geographical inequalities.

**Implications of all the available evidence:** National lockdowns are an important tool in the armoury of pandemic control, but their timing and duration must be carefully decided and be locally specific. Because case rate inequalities were already present before lockdown in England, movement restrictions served to further increase them.

**Summary Box:** 

**Section 1: What is already known on this subject:** Geographical inequalities in COVID-19 case rates have been noted worldwide, and in England. However, how these inequalities were affected by policy responses – such as national lockdowns - has yet to be investigated.

**Section 2: What this study adds:** We examined geographical inequalities in COVID-19 case rates by deprivation during the first English lock down (March – July, 2020). We find that cases were first reported in the less deprived areas of England, but this pattern quickly reversed and large excesses of cases occurred in the most deprived areas during the first national lockdown. Case rates in more deprived areas also rose more sharply, peaked higher, and then dropped faster than in less deprived areas. Inequality in cumulative case rates grew over the lockdown, increasing inequalities in disease burden.

## Introduction

Inequalities in the burden of COVID-19 have been demonstrated globally^1^. These inequalities exist by age, sex, ethnicity, socio-economic status and community deprivation (1–9). Most of this work has been descriptive and focused on comparing total case rates over set time phases^1^. There has been no analysis to date of how these inequalities in cases arose, changed overtime and were shaped by national mitigation strategies (lock downs). Understanding how these inequalities first emerged at the start of the COVID-19 pandemic in 2020 – and the impact of national policy responses (lock downs) - could help uncover important modifiable drivers which could be leveraged to prevent worsening of inequalities in future waves, or future pandemics. The first stage of such work is identifying the inequalities and how they arose. Deprivation has been linked to COVID-19 case rates in England previously^11^, but this paper provides the first analysis of the evolution of area-level inequalities in COVID-19 by deprivation levels in the first wave of the pandemic (January to July 2020) in England – with a focus on the effects of the first national lockdown (March – July 2020).

Many countries employed national ‘lockdown’ orders in the early stages of the pandemic, in an attempt to curtail viral transmission and avoid health system collapse^1^. In the UK, the first such lockdown was announced on the 23^rd^ March 2020, and began to be released on the 1^st^ June 2020. In keeping with many other European countries, this was characterised by a 12 week ‘stay at home’ order (SI 350) - whereby people could go outside only for certain “very limited purposes”. Face-to-face education was suspended and many workplaces closed, with employees ordered to ‘work from home’ where possible. Most office based workers were able to work from home, but key workers in factories, transport, food retail and health care continued to work as usual. Many other staff were furloughed as businesses ceased trading-particularly in the hospitality, travel and retail sectors^12^. As national cases, hospitalisations and deaths started to fall the lockdown was gradually released over several months - culminating in the so-called ‘Super Saturday’ on 4^th^ July 2020 when many businesses (including hospitality) reopened – albeit with strict social distancing rules^13^. Such lockdowns are an important and useful tactic in the national response, but the rationale for their timings are rarely transparent, and their impacts may not be experienced equally by different areas given geographical inequalities in the progression of the pandemic. We therefore explore the impact of the first English national lock down on inequalities in COVID-19 cases.

Clouston *et al*. (2021) leveraged the theories of Fundamental Cause Theory (FCT, ^14^) and Stages of Disease (SOD, ^15^) to test the hypotheses that COVID-19 cases would initially occur in less deprived areas, would then spread to more deprived communities, and the greater overall burden of disease would be experienced by the most deprived^2^. Their hypotheses were supported by the evolution of the US COVID-19 epidemic^2^. England also exhibits strong area-level socio-economic inequalities; thus we would expect similar case patterning to occur in England^16^.

It has been noted previously that when national epidemic dynamics are used for local decisions, they can mask important sub-national variation in disease spread, thus mitigation strategies that rely solely on the national data could inadvertently worsen inequalities^17^. Previous descriptive studies of inequalities in COVID-19 cases have tended to focus on cumulative measures over set timespans, without documenting the disparities in evolution of case rates^5,8,18^. An understanding of how national mitigation strategies affect geographical inequalities in COVID-19 cases would help inform policies targeted at minimising viral spread whilst preventing the widening (or even actively decreasing) health inequalities.

In this paper, we interrogate COVID-19 cases data from the first wave of the pandemic in England to identify geographical inequalities in the evolution of local epidemics, and to describe the effects of the first national lockdown on these inequalities.

## Methods

Weekly counts of COVID-19 cases for the period 2020-03-15 to 2020-07-04 per Middle Super Output Area (MSOA) for England were downloaded from GOV.UK and converted to rates per 100,000 persons using Office for National Statistics (ONS) mid-2019 population estimates. MSOAs are small-area geographies used in Census data^19^. They are comprised of Lower Super Output Areas (LSOAs). MSOAs represent a minimum population of 5000 and the mean population per MSOA is 7200 persons. There are 6,791 MSOAs and 32,844 LSOAs in England. MSOAs^19^ with no data preceding ‘Super Saturday’ (2020-07-04) or with fewer than four weeks of data were excluded (n=2377). The Isles of Scilly (E02006781) and City of London (E02006781) were excluded due to systematic differences in case reporting, leaving 4412 MSOAs for analysis.

Rank of Index of Multiple Deprivation (IMD) for all LSOAs in England was obtained from the ONS, and the mean rank was calculated for each MSOA. The mean IMD ranks were divided into deciles, where decile 1 contained the most deprived 10% of MSOAs in England. IMD is the standard measure of area-level deprivation in England. It produces a ranking of areas in England based on relative local scores for: income, employment, health, education, crime, access to services and living environment^20^.

Case rates per MSOA were converted to 3-week rolling means, and metrics characterising each MSOA’s first wave of epidemic (up to 2020-07-04) were calculated, including; peak case rate, speed of increase from 25% of peak rate to peak, speed of descent from peak to 50% of peak rate, total cumulative case rate over the first wave, and cumulative case rate up until the announcement of the first UK national lockdown (2020-03-23). Metrics were associated with IMD using simple linear or logistic regression models where appropriate. Similarly, cases for all MSOAs within each decile of IMD were summed, and comparative metrics of each decile’s first wave were compared.

A sensitivity analysis was conducted on daily cases data at Local Authority (LA) level (n=309). These data were acquired from GOV.UK and extended further back in time than the MSOA-level data, thus capturing more of the earliest COVID-19 case information. Results from analyses of LA data were similar to those of MSOA data.

All analyses were conducted in R statistical software version 3.6.2.

### Patient and Public Involvement

Our public involvement panel inputted into project design and considered the research topic to be of contemporary importance and value. The data used do not require patient permissions for use and are publicly available.

## Results

Of the 4412 MSOAs studied, in 1773 (40%) their earliest recorded case rate was their highest, i.e. the case rate peaked earlier than these data started being recorded. A simple logistic regression using an ‘early peak’ as the dependent binary variable showed that those MSOAs that peaked early were more likely to have a higher mean average rank of IMD (i.e. to be less deprived. IMD coefficient 1.001e-05, *p*-value 0.008).

Excluding the aforementioned MSOAs that peaked before data recording began, the peak case rate was higher in areas with low IMD (Figure 1). Each unit increase in average rank of IMD was associated with a very small (0.001 cases per 100,000 people) but statistically significant reduction in the peak case rate (Figure 2). There was no significant difference in the speed of increase or decrease in case rates across IMD at MSOA level.

**Figure 1.**
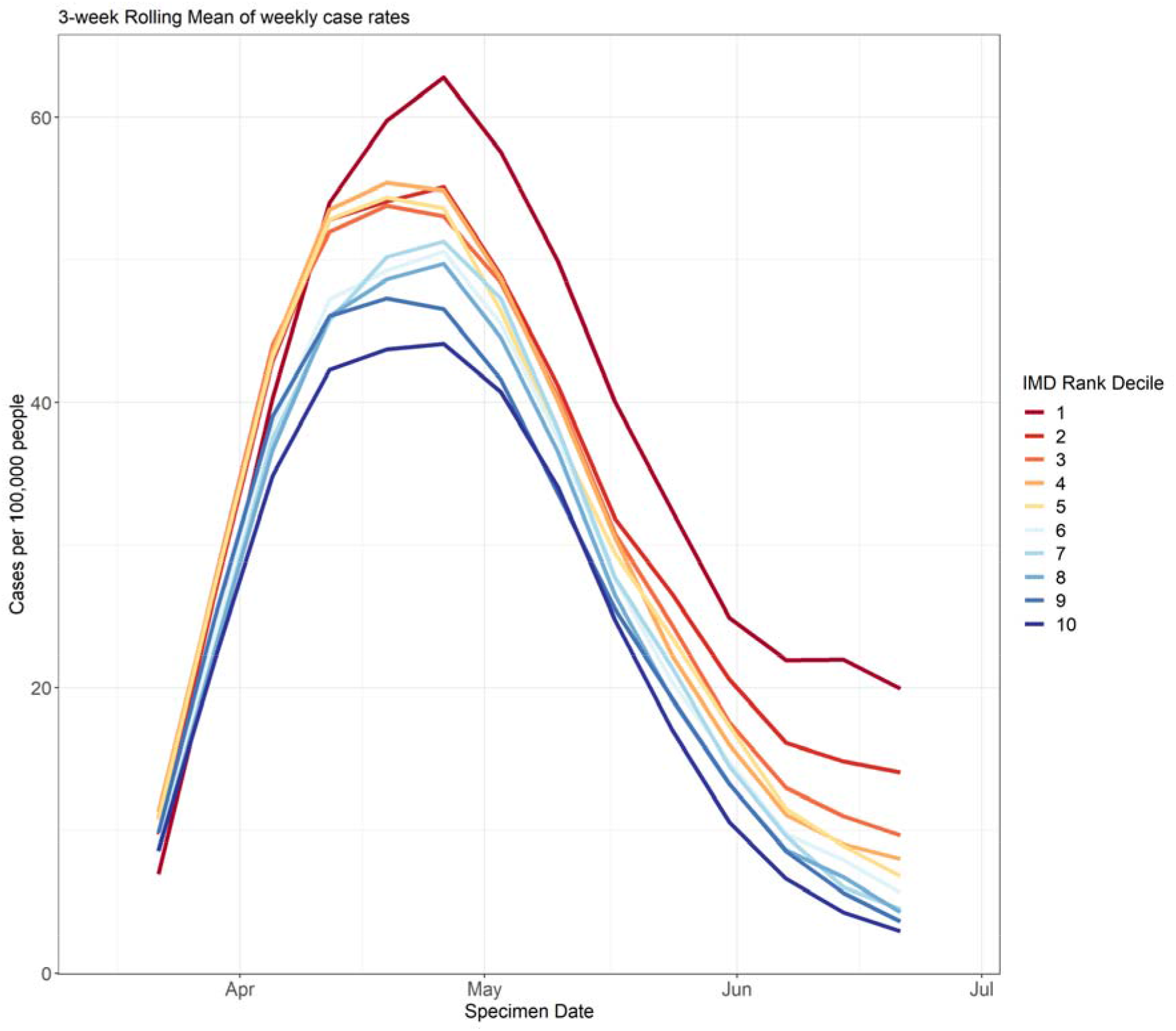
Mean daily case rates per 100,000 persons for MSOAs (n=4412) in each decile of rank of IMD (Index of Multiple Deprivation) across the first wave of the COVID-19 pandemic in England. Decile 1 is the most deprived group.

**Figure 2.**
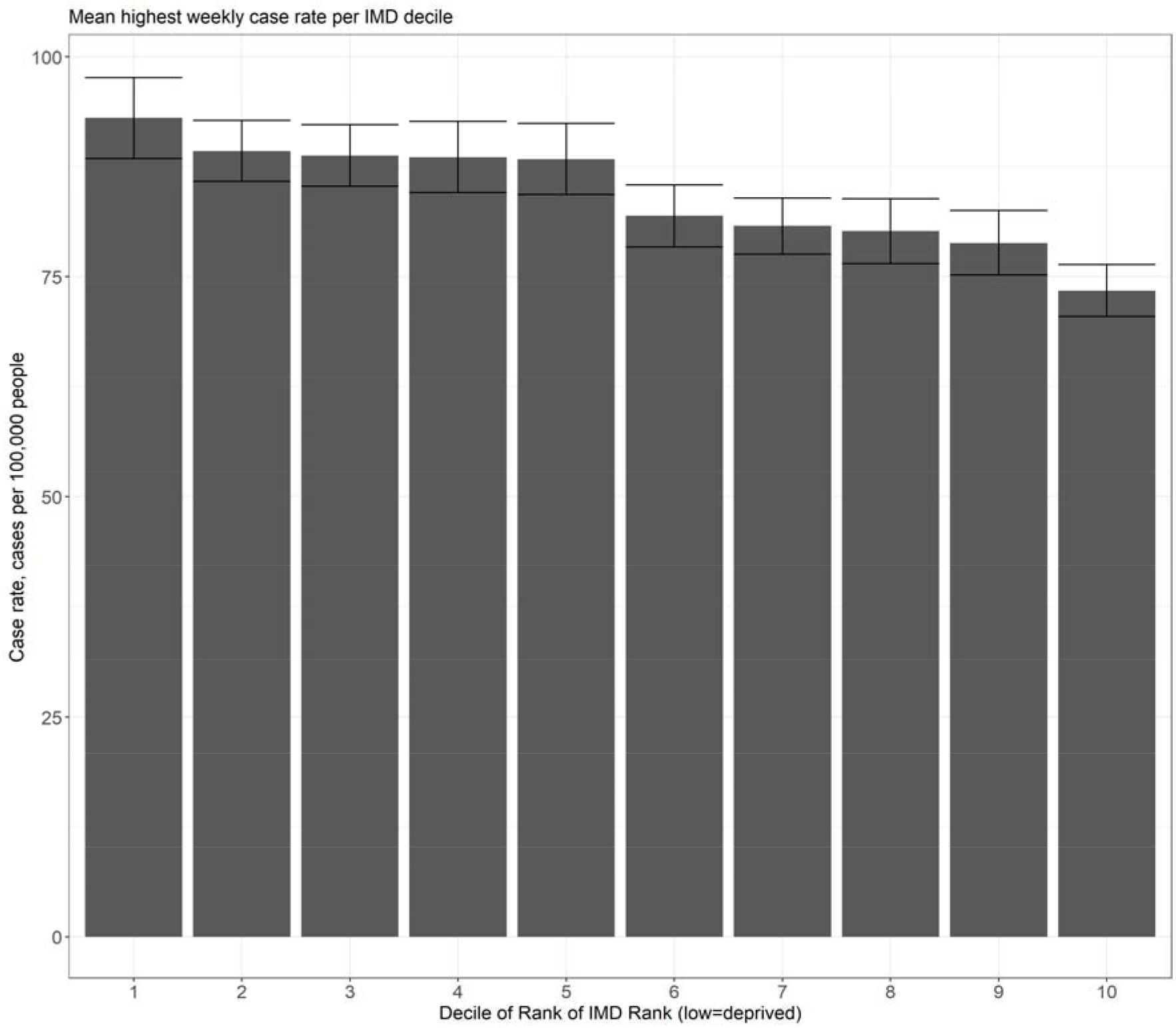
Mean peak case rate per decile of rank of IMD (Index of Multiple Deprivation) during the first wave of the COVID-19 pandemic in 4412 MSOAs (Middle Super Output Areas) in England.

Total cumulative case rates across the first wave were significantly lower in less deprived MSOAs compared to more deprived MSOAs (Figure 3).

**Figure 3.**
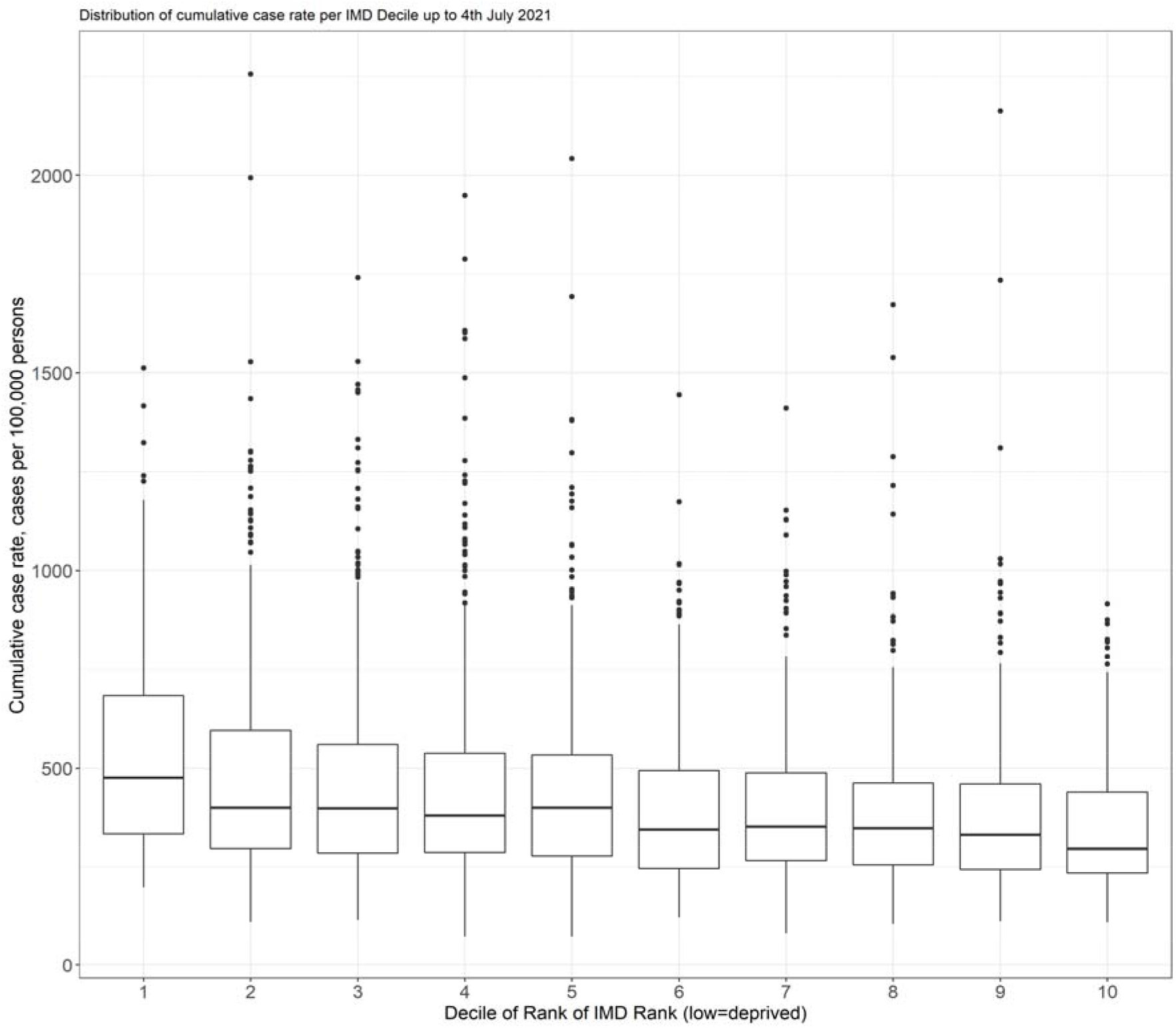
Boxplot of total cumulative case rates for all MSOAs (Middle Super Output Areas, n=4412) in each decile of rank of IMD (Index of Multiple Deprivation) over the first wave of COVID-19 pandemic in England.

When comparing the most deprived 20% of MSOAs in England (IMD deciles 1 and 2) with the least deprived 20% (IMD deciles 9 and 10), the ratio of cumulative case rates in most versus least deprived increased steadily across the first wave, showing that inequality in case rates grew over this period (Figure 4).

**Figure 4.**
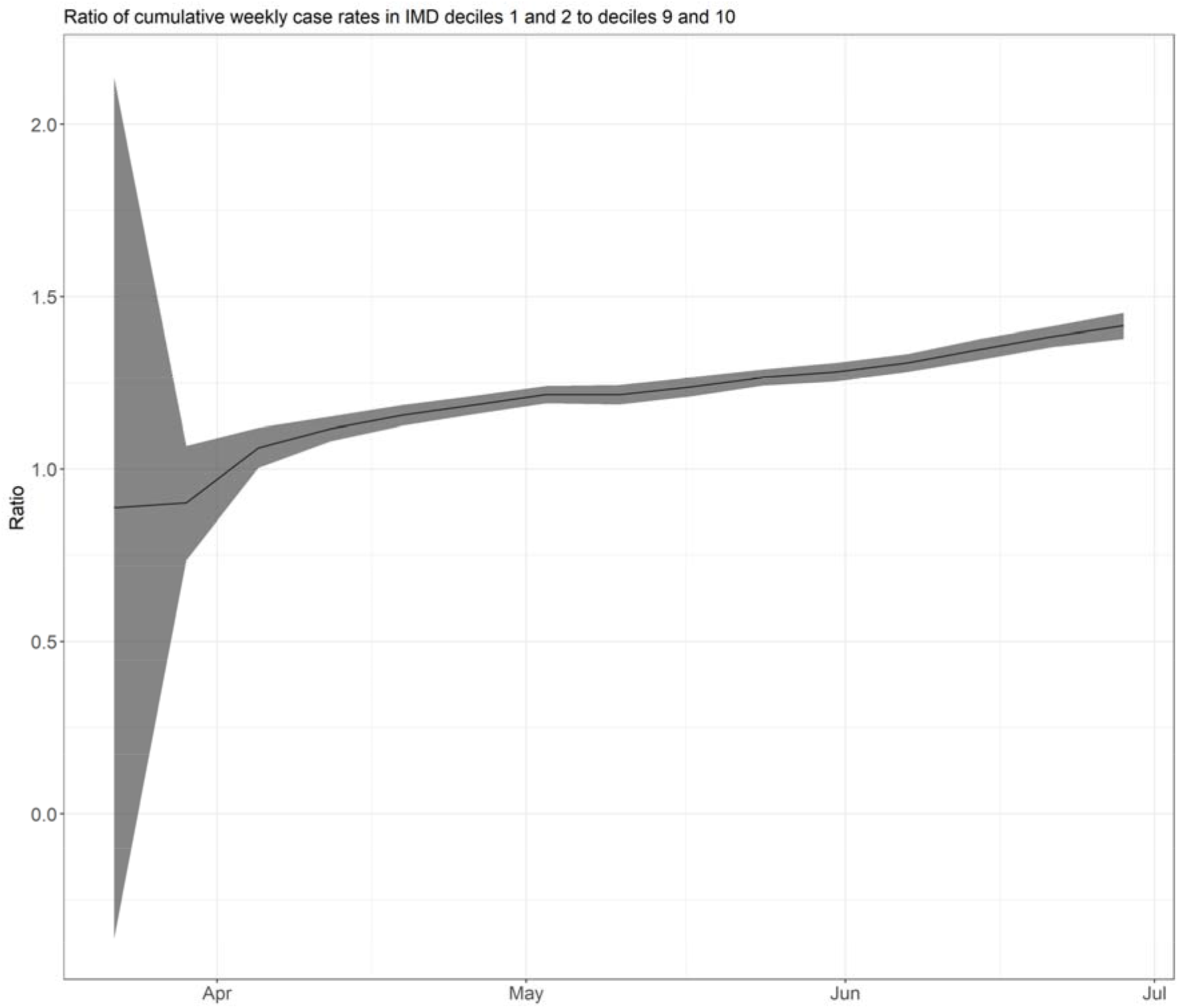
Ratio of least deprived/most deprived 20% of MSOAs (Middle Super Output Areas) cumulative case rates over the first wave of the COVID-19 pandemic in England.

In each MSOA, the weekly case rate up until the first UK national lockdown was announced (2020-03-23) was summed. The mean summed rate per IMD decile was then calculated, and the most and least deprived 20% of MSOAs were compared. Up until lockdown, the most deprived 20% of MSOAs recorded, on average, 42.4 cases per 100,000 persons, whereas the least deprived 20% (IMD deciles 9 and 10) recorded an average of 48.2 cases per 100,000 persons, thus the less deprived areas had accrued 14% more cases before lockdown began. At the end of the lock down period, the most deprived 20% had recorded on average 504.5 cases per 100,000, compared to the least deprived 20% which recorded 382.5 cases per 100,000 persons, i.e. cases were over 30% higher in the most deprived 20% of areas than in the least deprived 20%, thus inequality in case rates grew under the lockdown.

### Sensitivity Analysis: Local Authority level

The above metrics were calculated using similar methods for 309 (of 311 for which daily COVID-19 case numbers were available from GOV.UK for the period in question, and additionally excluding London and the Isles of Scilly) local authorities (LAs) across England. This sensitivity analysis was conducted to assess whether limitations of MSOA-level data (only available weekly and starting from a later date) were important. All comparable MSOA-level results were similar to those from LA-level analyses. In LA-level analyses, more deprived areas saw faster increases and decreases in case rates compared to less deprived areas. As local authority data were recorded earlier than MSOA-level data, we used these to determine whether a relationship existed between IMD and the timing of the first recorded COVID-19 cases per LA. We found that more deprived LAs began recording cases earlier than less deprived LAs (see Supplement).

## Discussion

In keeping with previous work, our study has found high inequalities in COVID-19 case rates by deprivation across England^1,20^. We have also found new evidence that the significant inequalities in the evolution of local COVID-19 epidemics have led to large differences in cumulative case numbers across England. Further, we have shown that these inequalities arose due to systematic differences in when the first cases were recorded, how quickly they grew, and to what peak level. The effect of the first UK national lockdown was to increase the relative excess of cases in deprived areas, likely due to a combination of factors – discussed below. We used data from two levels of geography to assess our hypotheses, one of which covered a large number of small areas, but suffered from a restricted time window and reduced regularity of data reporting, and another with a smaller number of larger areas which began earlier and was available as daily counts. We thereby leveraged the benefits of both datasets and provide sensitivity analyses.

Seeding of the COVID-19 virus into England probably occurred due to more affluent holidaymakers returning from Northern Italy^21^. From there, it spread within less deprived communities first, before viral spread in those areas was inhibited by lockdown. During the lockdown, residents of more deprived areas suffered from higher exposure levels and higher rates of transmission^6^. For example, were less able to fully isolate, due in part to more overcrowded housing, low/no sickness payments, lack of access to outdoor space, greater proportions of key-workers and people in public-facing roles^6^. Therefore, under movement restrictions, the virus was better able to spread within deprived communities. Throughout the lockdown, the case rate inequality grew. Our findings align with the fundamental causes theory proposed by Clouston *et al*. (2021) on stages of disease transmission and the generation of health inequalities^2^. However, our recent analyses of geographical inequalities in mortality found that whilst death rates were higher across the first wave in more deprived areas, the lock down did reduce them^22^.

Reduced viral load across the country is an important aim that benefits all inhabitants, through a reduction in selection pressure on the virus, and reduced demand on healthcare. Lockdowns are an effective means of achieving this aim quickly, however their potential harms need to be carefully weighed, and their timings carefully considered – and to reflect both local and national need. Based on our analyses, extended lockdowns may drastically increase health inequalities in case rates, for a population whose risk of adverse outcomes upon infection are already greater than others^23^. The duration of lockdown needs to consider the prevailing case rates at the time of initiation. The higher case rates are, the longer a lockdown will need to be to bring them under ‘control’. Given that lockdowns increase case rate inequalities, minimising this disparity relies upon their timely initiation in local areas before rates increase too much.

Using two different data sources to examine case rates, one at a higher geographical level but with earlier, more frequent, recording, and another at a lower level but with more a restricted timespan, we are able to identify patterns in local epidemics that would be hidden from one or other dataset. We found significant relationships between the speed of increase and decrease of case rates with IMD in LA-level data, which were not apparent in MSOA-level data. This discrepancy is likely due to the differences in data structure and reporting frequency in each, where daily data allow for a larger set of timepoints to examine trends compared to weekly data.

## Strengths and Limitations

In this study we leveraged the benefits of two separate datasets of case counts to avoid issues of small sample size, infrequent data recording and restricted time windows. MSOA-level data allowed for a sample size of 4412 small areas, with populations likely to be more homogeneous than at larger geographies. However, these data were only captured weekly beginning in March, when cases had already begun to be recorded in England. We excluded many MSOAs from some analyses whose cases were already declining at the earliest dates captured. To better understand the earliest phase of wave 1 in England, we therefore used LA-level data, which dated back further. These data had a more restrictive sample size (n=309) and covered a much larger population per area, but benefitted from being recorded daily. Significant differences in speeds of growth and decline in case rates were not found in MSOA-level data, despite being apparent at LA-level, likely because of restricted numbers of data points per area (weekly data). In general, the analyses of each dataset were similar, and combining both we were able to mitigate some of the limitations of each. The ideal data for this work would be at LSOA level, daily, extending back to 2020-01-01, however such data are not available for research and would suffer heavily from censoring due to low numbers.

Case rates in the early pandemic were likely heavily influenced by the availability of testing, which was initially only available in hospitals. This will have led to underreporting of the true community case rate, however such underreporting was likely similar across all areas and would therefore not have meaningfully affected comparative analyses.

## Supporting information

We found that more deprived LAs began recording cases earlier than less deprived LAs (see Supplement).

## Data Availability

All data used are publicly freely available through the GOV.UK and the ONS. Code used in the analyses is available upon request.

## Contributors

FM, CB and CW designed the study. CW completed all analyses with input from FM and CB. CW, VA, FM and CB all contributed to drafting the manuscript. The corresponding author attests that all listed authors meet authorship criteria and that no others meeting the criteria have been omitted. CW is guarantor of the analysis.

## Data sharing statement

All data used are publicly available at no cost.

## References

1 Bambra C, Lynch J, Smith KE. Unequal Pandemic: COVID-19 and Health Inequalities. Bristol: Policy Press, 2021 https://policy.bristoluniversitypress.co.uk/the-unequal-pandemic.

2 Clouston SAP, Natale G, Link BG. Socioeconomic inequalities in the spread of coronavirus-19 in the United States: A examination of the emergence of social inequalities. Social Science and Medicine 2021; 268: 113554.

3 Plümper T, Neumayer E. The pandemic predominantly hits poor neighbourhoodsã SARS-CoV-2 infections and COVID-19 fatalities in German districts. European journal of public health 2020; 30: 1176–80.

4 NHSA. COVID-19 and the Northern Powerhouse: Tackling inequalities for UK health and productivity. 2020.

5 Baena-Diéz JM, Barroso M, Cordeiro-Coelho SI, Diáz JL, Grau M. Impact of COVID-19 outbreak by income: Hitting hardest the most deprived. Journal of Public Health (United Kingdom) 2020; 42: 698–703.

6 Bambra C, Riordan R, Ford J, Matthews F. The COVID-19 pandemic and health inequalities. Journal of Epidemiology and Community Health 2020; 74: 964–8.

7 Public Health England. Disparities in the risk and outcomes of COVID-19. 2020; : 89.

8 Chen JT, Krieger N. Revealing the unequal burden of COVID-19 by income, race/ethnicity, and household crowding: US county versus zip code analyses. Journal of Public Health Management and Practice 2021; 27: S46–56.

9 Nicodemo C, Barzin S, Cavalli N, et al. Measuring geographical disparities in England at the time of COVID-19: results using a composite indicator of population vulnerability. BMJ open 2020; 10: e039749.

10 Sun Y, Hu X, Xie J. Spatial inequalities of COVID-19 mortality rate in relation to socioeconomic and environmental factors across England. Science of the Total Environment 2021; 758. DOI:10.1016/j.scitotenv.2020.143595.

11 Morrissey K, Spooner F, Salter J, Shaddick G. Area level deprivation and monthly COVID-19 cases: The impact of government policy in England. Social Science & Medicine 2021; 289: 114413.

12 HM Government. The Health Protection (Coronavirus, Restrictions) (England) Regulations 2020. SI 350. 2020. https://www.legislation.gov.uk/uksi/2020/350/contents/made (accessed Feb 23, 2021).

13 HM Government. The Health Protection (Coronavirus, Restrictions) (No. 2) (England) Regulations 2020. SI2020/684. 2020. https://www.legislation.gov.uk/uksi/2020/684/contents/made (accessed Feb 23, 2021).

14 Phelan JC, Link BG, Tehranifar P. Social Conditions as Fundamental Causes of Health Inequalities: Theory, Evidence, and Policy Implications. Journal of Health and Social Behavior 2010; 51: S28–40.

15 Clouston SAP, Rubin MS, Phelan JC, Link BG. A Social History of Disease: Contextualizing the Rise and Fall of Social Inequalities in Cause-Specific Mortality. Demography 2016; 53: 1631–56.

16 Bambra C, Barr B, Milne E. North and South: Addressing the English health divide. Journal of Public Health (United Kingdom) 2014; 36: 183–6.

17 Salvatore M, Basu D, Ray D, et al. Comprehensive public health evaluation of lockdown as a non-pharmaceutical intervention on COVID-19 spread in India: National trends masking state-level variations. BMJ Open 2020; 10. DOI:10.1136/bmjopen-2020-041778.

18 Niedzwiedz CL, O’Donnell CA, Jani BD, et al. Ethnic and socioeconomic differences in SARS-CoV2 infection in the UK Biobank cohort study. medRxiv 2020; : 2020.04.22.20075663.

19 Office for National Statistics. Census Geography. 2012. https://www.ons.gov.uk/methodology/geography/ukgeographies/censusgeography#output-area-oa.

20 Ministry of Housing C& LG. English indices of deprivation. 2020. https://www.gov.uk/government/statistics/english-indices-of-deprivation-2019.

21 Correa-Martínez CL, Kampmeier S, Kümpers P, et al. A pandemic in times of global tourism: Superspreading and exportation of COVID-19 cases from a ski area in Austria. Journal of Clinical Microbiology 2020; 58: 19–21.

22 Welsh CE, Albani V, Matthews FE, Bambra C. Geographical inequalities in the evolution of the COVID-19 pandemic: An ecological study of inequalities in mortality in the first wave and the effects of the first national lockdown in England. medRxiv 2021. DOI:https://doi.org/10.1101/2021.10.23.21265415.

23 Tian W, Jiang W, Yao J, et al. Predictors of mortality in hospitalized COVID-19 patients: A systematic review and meta-analysis. Journal of Medical Virology 2020; : 1–9.

