## Supplementary material for "The effects of the first national lockdown in England on geographical inequalities in the evolution of COVID-19 case rates: An ecological study": We found that more deprived LAs began recording cases earlier than less deprived LAs (see Supplement).

The first week in which LAs began recording cases ranged from 2020-01-30 to 2020-03-31, with most (79%) beginning in March. For each week in March 2020 (weeks 9 to 14), the proportion of cases that occurred in LAs of each IMD decile is shown in Figure S1. There is a visual trend towards less deprived LAs beginning their epidemics earlier than more deprived LAs.

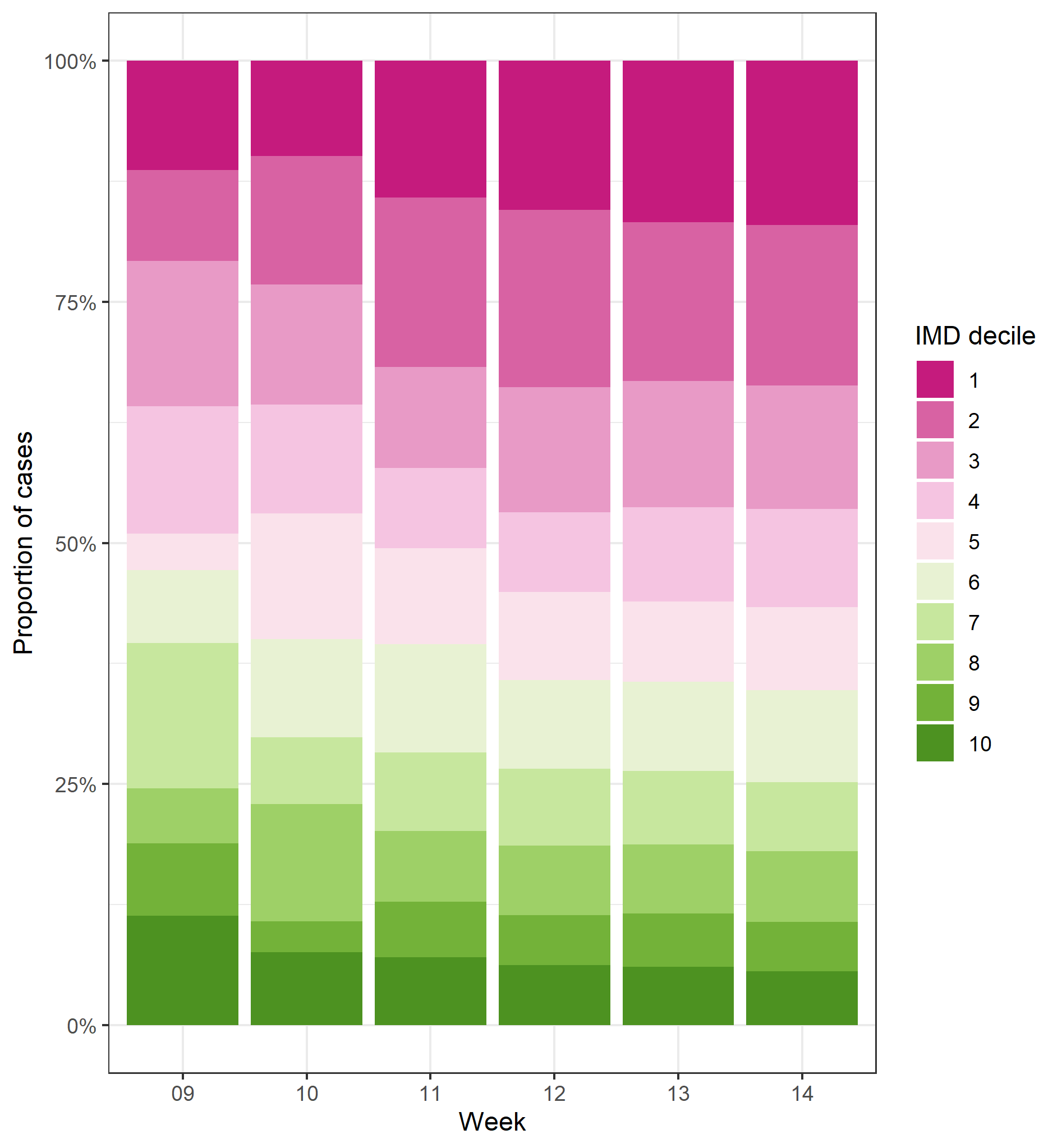

**Figure S1.** Breakdown by IMD decile of the local authorities that started recording COVID-19 cases in each week of March 2020 (n=264).

The speed of increase in case rates was greater in more deprived LAs (1 unit higher IMD rank associated with -0.003 cases per week gradient of increase, *p-*value <0.001), and their peak case rates were higher than less deprived areas (1 unit higher IMD rank associated with -0.07 cases per 100,000 at peak, *p-*value <0.001). The speed of descent in case rates from the peak to 50% of peak was faster in more deprived LAs (1 unit higher IMD rank associated with -0.001 cases per 100,000 persons per week, *p*-value <0.001).

At the LA level, using all available data up to the point of the first lockdown, LAs in deciles 9 and 10 (least deprived 20% of LAs in England) had recorded fewer cases than the most deprived 20% (mean of 13.7 versus 15.9 cases per 100,000 persons). Over the whole of the first wave, this gap had increased, with the total cumulative case rate in the least deprived LAs amounting to 67% of that of the most deprived (338.5 versus 508.0 cases per 100,000 persons). As with the MSOA-level analyses, the period of lockdown appeared to favour less deprived LAs compared to the more deprived, in terms of cumulative case numbers.
